# Time courses of COVID-19 infection and local variation in socioeconomic and health disparities in England

**DOI:** 10.1101/2020.05.29.20116921

**Authors:** Shelley H. Liu, Bian Liu, Yan Li, Agnes Norbury

## Abstract

**Objective:** To identify factors associated with local variation in the time course of COVID-19 case burden in England.

**Methods:** We analyzed laboratory-confirmed COVID-19 case data for 150 upper tier local authorities, from the period from January 30 to May 6, 2020, as reported by Public Health England. Using methods suitable for time-series data, we identified clusters of local authorities with distinct trajectories of daily cases, after adjusting for population size. We then tested for differences in sociodemographic, economic, and health disparity factors between these clusters.

**Results:** Two clusters of local authorities were identified: a higher case trajectory that rose faster over time to reach higher peak infection levels, and a lower case trajectory cluster that emerged more slowly, and had a lower peak. The higher case trajectory cluster (79 local authorities) had higher population density (p<0.001), higher proportion of Black and Asian residents (p=0.03; p=0.02), higher multiple deprivation scores (p<0.001), a lower proportions of older adults (p=0.005), and higher preventable mortality rates (p=0.03). Local authorities with higher proportions of Black residents were more likely to belong to the high case trajectory cluster, even after adjusting for population density, deprivation, proportion of older adults and preventable mortality (p=0.04).

**Conclusion:** Areas belonging to the trajectory with significantly higher COVID-19 case burden were more deprived, and had higher proportions of ethnic minority residents. A higher proportion of Black residents in regions belonging to the high trajectory cluster was not fully explained by differences in population density, deprivation, and other overall health disparities between the clusters.

**What is already known on this subject?:** Emerging evidence suggests that the burden of COVID-19 infection is falling unequally across England, with provisional data suggesting higher overall infection and mortality rates for Black, Asian, and mixed race/ethnicity individuals.

**What does this study add?:** We found that regions with greater socioeconomic deprivation and poorer population health measures showed a faster rise in COVID-19 cases, and reached higher peak case levels. Areas with a higher proportion of Black residents were more likely to show this kind of time course, even after adjusting for multiple co-occurring factors, including population density. This finding merits further investigation in terms of the intersecting vulnerability factors Black and other minority ethnic individuals face in England (e.g. proportion of people working in service and caring roles, and the role of structural discrimination), and has implications for the ongoing allocation of public health resources, in order to better mitigate such inequalities.

## Introduction

England currently has amongst the highest worldwide recorded cases of the novel coronavirus (COVID-19), with 243,303 cases as of May 17, 2020 [1]. However, increasing evidence suggests this burden disproportionately affects certain groups. Provisional data from the Office for National Statistics has indicated significantly increased COVID-19 mortality estimates for Black, Asian, and mixed race/ethnicity individuals, even after indirect adjustment for age, broad geographical region, and measures of self-reported health and disability [2,3]. There are also disparities in COVID-19 mortality across geographic regions. Based on COVID-19 deaths occurring in March and early April, the age-standardized mortality rate was twice as high in the most deprived areas of England, compared with the least deprived [4]. Importantly, a recent study assessing hypothetical vulnerability to respiratory pathogen pandemics – calculated in terms of likely demand characteristics versus supply-side differences in existing healthcare provision – found greater vulnerability in regions of England with higher economic deprivation levels [5].

Here, we sought to identify whether local-level variation in *trajectories* (changes over time) in confirmed COVID-19 cases were related to variation in socioeconomic deprivation and self-reported racial/ethnic identity. We also assessed whether time courses of caseloads differed according to important population health measures – including healthy life expectancy, preventable mortality, and proportion of individuals with chronic health conditions that may increase susceptibility to COVID-19 morbidity and mortality. The ability of different areas to control COVID-19 infection rates over time may provide a more sensitive readout of vulnerability than cumulative case or mortality data, as this time course will depend on a combination of sociodemographic variables and local capacity to enact public health measures that help limit disease spread [5]. Critically, we examined whether differences in demographic measures between trajectories remained significant after adjusting for covariance with other factors that may affect COVID-19 caseload (population density, proportion of older adults, relative deprivation, and preventable mortality rates) [6–8].

## Methods

Data on daily laboratory-confirmed COVID-19 cases for the period spanning January 30 to May 6, 2020 were accessed via Public Health England [9]. These case counts are reported based on the date the specimen was taken from the patient, not the date the lab carried out testing and reported results to Public Health England. Case data were extracted for upper tier local authorities (henceforth referred to as local authorities), a level of analysis which was chosen in order to balance geographical resolution against availability of up-to-date clinical and sociodemographic data. Case counts were adjusted for local authority population size, using population estimates provided by the Office for National Statistics (ONS), as of April 2019 [10].

We identified clusters, or homogenous groups, of local authorities with distinct COVID-19 case trajectories. Clusters were identified using a partition around mediods approach [11], a clustering analysis for time-series data. A dynamic time warping similarity metric was used to capture nonlinear similarity between two time-series based on their shape. This metric was chosen because it is has been shown in the literature to be optimal for clustering of time series data. Unlike the Euclidean distance which is commonly used for cluster analysis of non-time series data, the dynamic time warping distance is not sensitive to time shifts, which is helpful as some local authorities may have had earlier cases than others. The clustering algorithm partitions local authorities in order to maximize intra-cluster similarities in case trajectories, and maximize inter-cluster differences in case trajectories. In order to identify the optimal number of clusters, 2-, 3-, 4-, and 5-cluster models were compared using five established cluster validity indices (Silhouette, Score function, Davies-Bouldin, modified Davies-Bouldin, and COP) [12].

Sociodemographic, economic deprivation and public health data for each local authority were compiled from Public Health England Fingertips repository using FingertipsR [13]. Specifically, we extracted the most recent data on ethnicity (proportions of Black or Black British ethnic group, Asian or Asian British ethnic group, Mixed/multiple ethnic group and White residents); older adults (proportion of adults aged 65 or older), index of multiple deprivation (IMD, a composite measure of deprivation calculated across multiple domains, including income, employment, education, health, crime, barriers to housing and services, and living environment: [14]); employment deprivation (proportion of working-age population involuntarily excluded from the labor market, calculated from claimants of various out-of-work social security allowances: [14]), healthy life expectancy (average number of years a person would expect to live in good health based on contemporary mortality rates and prevalence of self-reported good health); preventable mortality (age-standardized mortality rate from causes considered preventable, per 100,000 people); pre-existing chronic health conditions (proportion of registered patients with a general practitioner (GP)-recorded diagnosis of coronary heart disease, diabetes, hypertension or obesity); self-reported physical activity (proportion of adults completing at least 150 minutes of moderate-intensity physical activity per week); and nursing home admissions (permanent admissions to residential and nursing care homes, per 100,000 people aged 65+). Population density was calculated as number of people per land area square kilometer, according to standard area measurements of local authorities available from the Office of National Statistics Open Geography portal [15].

Differences in sociodemographic, deprivation and public health measures between the clusters were tested using the nonparametric Kruskal Wallis test. We presented the median and interquartile range (IQR) of the measures in each cluster. In addition, separate logistic regression models were used to determine whether proportions of minority ethnicity groups and deprivation predicted cluster membership, after adjusting for potential confounding variables. Odds ratios (OR) and 95% confidence intervals (CI) for adjusted models are presented. All statistical analyses were carried out using R, version 3.6.1 (R Core Team, 2019).

## Results

Two clusters of local authorities were identified, based on daily population-adjusted COVID-19 laboratory confirmed case counts (Figure 1a). All five cluster validity indices supported the identification of two clusters. A cluster of 79 local authorities had significantly higher population-adjusted daily case trajectories. Members of this cluster tended to be located close to or contain major urban centers (including Manchester, Newcastle upon Tyne, Liverpool, Birmingham, Sheffield and London), however members of this cluster also included less densely populated areas (such as Cumbria, Northumberland, and Herefordshire; Figure 1b). A second cluster of 71 local authorities followed a lower daily case trajectory. These areas tended to be less densely populated, including the south-west region, but also included several London boroughs (Hillingdon, Richmond, Westminster, Camden, Islington, City of London, Tower Hamlets and Greenwich; Figure 1b, inset).

**Figure 1.**
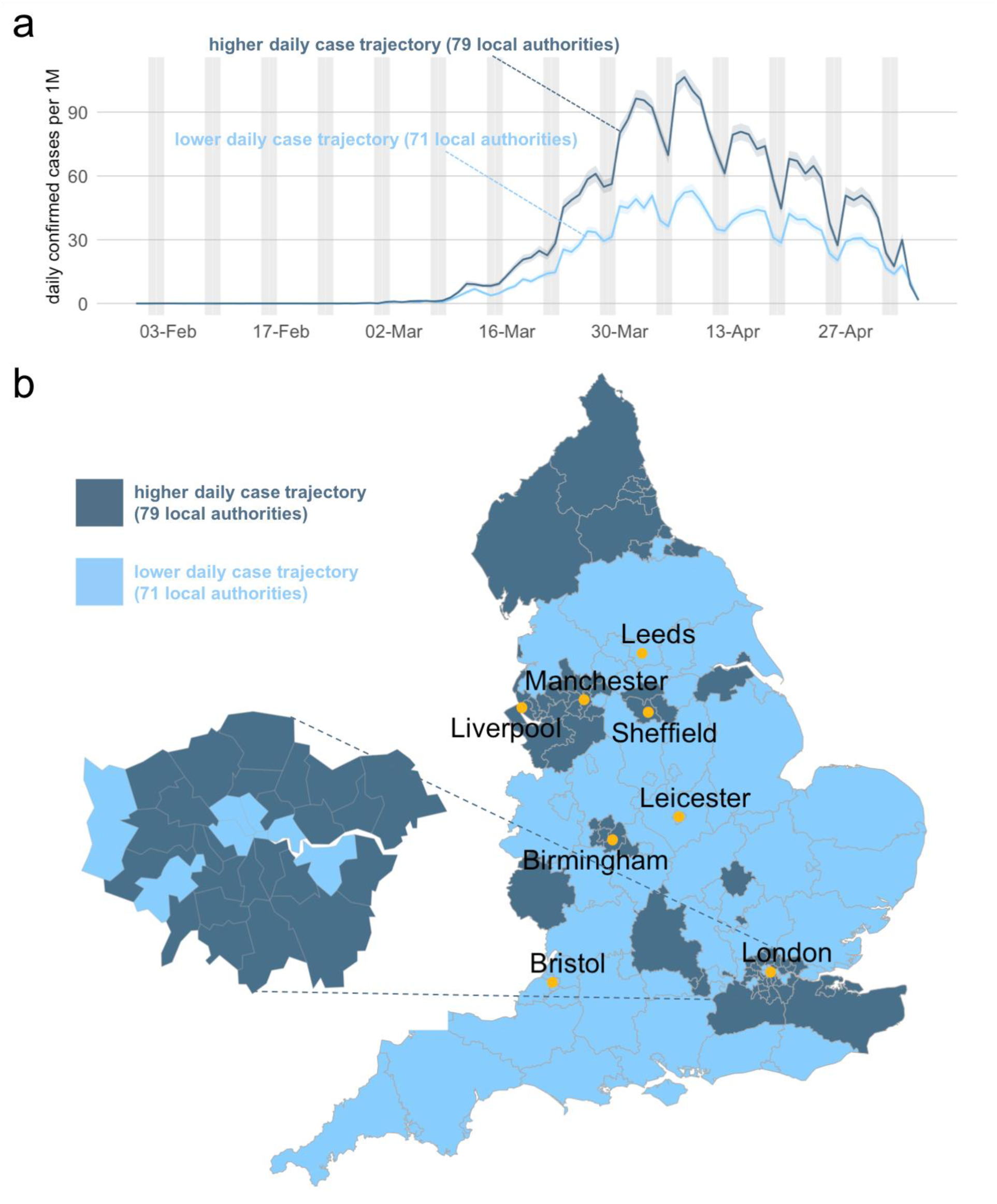
Cluster analysis of daily COVID-19 case data by local authority region. **a** Analysis of daily case data identified two trajectory clusters: a higher daily case trajectory (formed of 79 local authorities), and a lower case trajectory cluster (formed of 71 local authorities. Data plotted here represent the mean (standard error) of laboratory-confirmed COVID-19 cases each day, per 1 million (1M) population (data from Public Health England). Grey shading represents weekends (Saturdays and Sundays), when number of reported cases tends to be lower. **b** Cluster assignment, plotted for each region (unitary or upper tier local authorities). Boundary data are correct as of December 2019 (source: Office for National Statistics Open Geography portal). Inset shows a magnification of the Greater London area. The eight largest cities by population size are labeled.

Both clusters followed a similar temporal pattern, with case counts increasing until early April and then declining. However, as of May 6, 2020, the median cumulative number of cases per 1 million population in the high trajectory cluster was 2947.64 [IQR: 2543.85, 3350.20], as compared with 1717.96 [IQR: 1376.34, 1999.41] cases per 1 million population in the low trajectory cluster (*p*<0.001). Figure 1a shows that this was due to a faster rate of rise, and higher overall peak level in the higher case, compared to the lower case trajectory cluster.

Table 1 presents the socio-demographic and health factors in the two clusters. Local authorities in the high trajectory cluster had significantly higher proportions of Black and Black British ethnic group residents (*p*=0.026). Local authorities in the high trajectory cluster had a median of 2.36% [IQR: 0.49%, 8.55%] Black residents, compared with a median of 0.89% [IQR: 0.57%, 2.14%] Black residents in the low trajectory cluster group. There was also higher proportions of Asian and Asian British ethnic group residents in the high trajectory cluster, with a median of 7.41% [IQR: 2.33%, 15.57%] as compared with a median of 3.71% [IQR: 1.80%, 8.28%] in the low trajectory cluster group. Local authorities in the high trajectory cluster group had significantly lower proportions of white residents (*p*=0.011), while they did not differ on the proportions of Mixed/multiple ethnic group residents (*p*=0.101).

**Table 1.** Socio-demographic, health factors and burden of COVID-19. ^1^Black denotes Black or Black British ethnic group;^2^Asian denotes Asian or Asian British ethnic group;^3^IMD, Index of Multiple Deprivation, a composite deprivation score encompassing income, employment, education, health, crime, barriers to housing and services, and living environment;^4^Employment deprivation scale (proportion of the working-age population in an area involuntarily excluded from the labour market, due to factors such as unemployment, sickness, disability or carer responsibilities) represents the most recently available data, from 2015.^5^Population density is defined as number of persons per land area square kilometer. ^6^Healthy life expectancy (average number of years a person would expect to live in good health based on contemporary mortality rates and prevalence of self-reported good health: M, for males; F, for females);^7^Preventable mortality represents the age-standardized mortality rate from causes considered preventable per 100,000 population for 2016–2018 (M, for males; F, for females). ^8^Chronic conditions: percentage of registered patients with a general practitioner (GP)-recorded diagnosis of coronary heart disease, diabetes, hypertension or obesity;^9^permanent admissions to residential and nursing care homes, per 100,000 people aged 65+.^10^Cumulative COVID-19 infections per 1 million (1M) population are those reported as of May 6, 2020. \**p*<0.05, \*\**p*<0.01; Kruskal Wallis tests. Data provided by Public Health England.

|  | High trajectory cluster<br>(n=79) | Low trajectory cluster<br>(n=71) |  |
| --- | --- | --- | --- |
|  | Median (interquartile<br>range) | Median (interquartile<br>range) | p-value |
| **Racial/ethnic composition and older adults** |  |  |  |
| <sup>1</sup>Black (%) | 2.36 [0.49, 8.55] | 0.89 [0.57, 2.14] | 0.026 |
| <sup>2</sup>Asian (%) | 7.41 [2.33, 15.57] | 3.71 [1.80, 8.28] | 0.016 |
| Mixed/Multiple ethnic<br>group (%) | 2.08 [1.07, 4.17] | 1.57 [1.17, 2.48] | 0.101 |
| White (%) | 85.49 [65.75, 95.97] | 92.66 [85.92, 95.74] | 0.011 |
| 65+ Male (%) | 15.16 [11.84, 18.20] | 17.18 [13.55, 20.11] | 0.005 |
| 65+ Female (%) | 17.98 [14.23, 20.95] | 20.06 [16.00, 22.88] | 0.004 |
| **Deprivation and population density** |  |  |  |
| <sup>3</sup>IMD score | 25.66 [19.24, 31.10] | 19.00 [14.76, 26.35] | <0.001 |
| <sup>3</sup>IMD decile | 4.00 [2.00, 7.00] | 7.00 [4.00, 9.00] | <0.001 |
| <sup>4</sup>Employment deprivation | 0.13 [0.11, 0.17] | 0.11 [0.08, 0.14] | <0.001 |
| <sup>5</sup>Population density<br>(persons per km<sup>2</sup>) | 2526 [1269, 4735] | 589 [322, 2546] | <0.001 |
| **Health factors** |  |  |  |
| <sup>6</sup>Healthy life expectancy<br>(M) | 61.42 [59.22, 64.68] | 63.15 [61.40, 65.66] | 0.009 |
| <sup>6</sup>Healthy life expectancy<br>(F) | 62.40 [59.17, 64.82] | 63.99 [60.99, 65.90] | 0.058 |
| <sup>7</sup>Preventable mortality per<br>100,000 (M) | 251.20 [218.40, 288.75] | 220.40 [195.30, 258.30] | 0.003 |
| <sup>7</sup>Preventable mortality per<br>100,000 (F) | 147.60 [124.20, 170.10] | 133.80 [117.40, 154.50] | 0.027 |
| <sup>8</sup>Diabetes (%) | 7.30 [6.53, 8.13] | 6.82 [6.12, 7.35] | 0.001 |
| <sup>8</sup>Overweight/Obese (%) | 64.04 [59.05, 67.15] | 62.66 [60.33, 65.44] | 0.196 |
| <sup>8</sup>Coronary Heart Disease<br>(%) | 3.12 [2.34, 3.85] | 3.12 [2.51, 3.69] | 0.934 |
| <sup>8</sup>Hypertension (%) | 13.80 [12.10, 15.70] | 14.27 [12.66, 15.97] | 0.363 |
| Physically active (%) | 65.60 [61.61, 68.74] | 68.12 [65.50, 70.97] | 0.001 |
| <sup>9</sup>Nursing home<br>admissions per 100,000 | 600.80 [452.20, 735.95] | 558.20 [460.60, 627.60] | 0.088 |
| <sup>10</sup>Cumulative COVID-19<br>infections per 1M | 2947.64 [2543.85,<br>3350.20] | 1717.96 [1376.34,<br>1999.41] | <0.001 |

Local authorities in the high trajectory cluster also had significantly lower proportions of older adult males and females aged 65+ (*p*=0.005; *p*=0.004). However, there were higher rates of admission into nursing homes among older adults aged 65+ in the high trajectory cluster (p=0.088).

There was significantly greater deprivation, quantified through both the multiple index of deprivation and employment deprivation, in the high case trajectory cluster (*p*<0.001; *p*<0.001). The high case trajectory cluster also had significantly greater population density (*p*<0.001). This was a median of 2526 [IQR: 1269, 4735] persons per land area square kilometer in the high case trajectory cluster, compared with 589 [IQR: 322, 2546] persons per land area square kilometer in the low case trajectory cluster.

The cluster of local authorities with a high daily case trajectory had significantly poorer health measures. It had higher preventable mortality per 100,000 persons for males and females (p=0.003; *p*=0.027), and had lower healthy life expectancy for males (*p*=0.009). Further, significantly lower proportion of adults were physically active (*p*=0.001), and there was higher prevalence of diabetes (*p*=0.001).

Table 2 presents results from logistic regression models. Because population density varies significantly between clusters, and high population density could facilitate the spread of an infectious disease, we adjusted for it in logistic regression models. In adjusted models, local authorities with higher proportions of Black residents were significantly more likely to belong to the high case trajectory cluster (OR 1.14 [95% CI: 1.02 – 1.29], *p*=0.026), and local authorities with higher proportions of White residents were significantly less likely to have high case trajectory membership (OR 0.97 [95% CI: 0.93, 0.99], *p*=0.041).

**Table 2.** Adjusted models of socio-demographic factors and burden of COVID-19. ^1^Black denotes Black or Black British ethnic group;^2^Asian denotes Asian or Asian British ethnic group.^3^IMD, Index of Multiple Deprivation, a composite deprivation score encompassing income, employment, education, health, crime, barriers to housing and services, and living environment.^4^Employment deprivation scale (proportion of the working-age population in an area involuntarily excluded from the labour market, due to factors such as unemployment, sickness, disability or carer responsibilities) represents the most recently available data, from 2015. Odds ratios and associated confidence intervals were calculated using logistic regression adjusted for different covariates. Data provided by Public Health England.

|  | **Odds ratio of**<br>**belonging to**<br>**high case**<br>**trajectory** | **95% CI** | **p-value** |
| --- | --- | --- | --- |
| **Models adjusted for population density** |  |  |  |
| <sup>1</sup>Black (%) | 1.14 | 1.02 – 1.29 | 0.026 |
| <sup>2</sup>Asian (%) | 1.03 | 0.99 – 1.08 | 0.134 |
| Mixed/Multiple ethnic<br>group (%) | 1.08 | 0.76 – 1.53 | 0.674 |
| White (%) | 0.97 | 0.93 – 0.99 | 0.041 |
| Older adults (%) | 0.92 | 0.81 – 1.03 | 0.140 |
| <sup>3</sup>IMD score | 1.08 | 1.03 – 1.14 | 0.001 |
| <sup>4</sup>Employment<br>deprivation | 1.19 | 1.09 – 1.31 | <0.001 |
| **Models adjusted for population density, % older adults, % Black, % Asian,**<br>**% Mixed/Multiple ethnic group** |  |  |  |
| <sup>3</sup>IMD score | 1.08 | 1.03 – 1.14 | 0.002 |
| <sup>4</sup>Employment<br>deprivation | 1.21 | 1.09 – 1.35 | <0.001 |
| **Models adjusted for population density, deprivation, % older adults and preventable**<br>**mortality** |  |  |  |
| <sup>1</sup>Black (%) | 1.20 | 1.03 – 1.45 | 0.035 |
| <sup>2</sup>Asian (%) | 1.02 | 0.97 – 1.08 | 0.465 |
| Mixed/Multiple ethnic<br>group (%) | 1.14 | 0.68 – 1.99 | 0.634 |
| White (%) | 0.95 | 0.90 – 1.00 | 0.083 |

Next, to identify whether higher proportion of minority ethnic groups is associated with higher probability of belonging to the high case trajectory, we also adjusted for additional covariates. Local authorities with higher proportions of Black residents were more likely to belong to the high case trajectory cluster (*p*=0.035), even after adjusting for population density, deprivation, proportion of older adults and preventable mortality. The adjusted odds ratio corresponding to a 1% increase in Black residents was 1.20 (95% CI: 1.03 – 1.45).

Higher index of multiple deprivation scores were also significantly associated with high case trajectory after adjusting for differences in population density, proportion older adults, and proportion of Black, Asian and Mixed/Multiple ethnic groups (OR = 1.08 [95% CI: 1.03 – 1.14], *p*=0.002). Further, in adjusted models, higher employment deprivation was significantly associated with high case trajectory (OR = 1.21 [95% CI: 1.09 – 1.35], *p*<0.001).

## Discussion

Our findings suggest that the burden of COVID-19 falls disproportionately on areas in England with higher deprivation, a higher proportion of Black and Asian residents, a higher prevalence of chronic diseases, more physical inactivity, and lower healthy life expectancy, and higher mortality from preventable causes.

Importantly, local differences in race/ethnicity and deprivation scores significantly predicted membership of the higher case trajectory cluster after adjusting for differences in population density – suggesting these effects are not simply the result of higher proportions of ethnic minority residents and higher deprivation levels in dense urban areas where COVID-19 is likely to be more prevalent [16].

The geographic disparities in case trajectories were in general consistent with the trend in COVID-19 mortality data, where the death toll in the most deprived areas of England and Wales were approximately twice that in the least deprived areas [17]. Often, populations living in deprivation also are co-living with a higher burden of chronic health conditions that elevate risk for COVID-19 morbidity and mortality, producing a compounded effect [18–20]. Similar health inequalities in COVID-19 outcomes have been reported elsewhere with regard to socioeconomic and demographic profiles. [21–24]. Our results are also aligned with estimates based on individual-level data from the UK Biobank, which showed a 2 to 4-fold higher risk of COVID-19 infection among Black, Asian, and minority ethnic (BAME) communities than their white counterparts, after adjusting for socioeconomic status, lifestyle, obesity, and comorbidities [25].

However, it is important to note that it is difficult to separate racial, socioeconomic, and health contributions when reporting COVID-19 disparities – and, crucially, that insufficient contextualization of findings may lead to misunderstandings that undermine the goal of eliminating health inequalities [26]. For example, recent analysis from the Office for National Statistics suggests that occupation may pay an important role in COVID-19 exposure and mortality – with workers in social care, public transport, and sales/retail all found to have increased death rates [27]. The increased exposure to other individuals and reduced ability to maintain physical distancing associated with these roles is likely to interact with findings stratified by ethnicity groups, as there are significantly greater proportions of BAME individuals employed in caring and customer service professions [28]. Further, there are demonstrable long-term health implications of structural inequality and discrimination with respect to race and ethnicity. Research has demonstrated that the cumulative effects of physiological responses to external stressors, especially racial discrimination, are linked to increased risk of chronic health conditions such as cardiovascular disease and diabetes (the ‘weathering’ hypothesis, [29,30]) – further exacerbating risk from COVID-19 [31,32]. Finally, migrant groups in the UK may have limited healthcare entitlements, and face additional barriers to healthcare access, including fear of the imposition of high charges and/or sharing of their data with immigration enforcement agencies [3,33].

Interestingly, areas with a high daily case trajectory had a significantly lower proportion of older adults (aged 65 or above), which preliminarily suggests that it may not be older population that is driving the higher burden of COVID-19 infection in these regions. However, as the healthy life expectancy of males is significantly lower in the high trajectory areas, and the admissions into nursing homes is higher in the high trajectory areas, it is possible that older adults in these areas may have poorer health; however, this warrants further study.

The burden of COVID-19 deaths has fallen disproportionately in major urban centers – particularly London, Liverpool, Birmingham and Manchester [4,17]. As expected, local authorities in these cities tended to belong to the high case trajectory cluster. However, we note that within London, some wealthier local authorities (e.g. Westminster, City of London) belonged to the lower case trajectory cluster. People with means may be more able to enact physical distancing, and have the ability to leave London, as 250,000 people were estimated to have left London in mid-March prior to the lockdown based on smartphone data [34].

This study had some limitations. There may be some local differences in availability and provision of COVID-19 testing, although this is not expected to be a significant limitation, as there is standardized government guidance for members of the public who suspect they have COVID-19, to contact NHS 111 or visit https://nhs.net, rather than contacting their GP. Further, there may be outflow and inflow of residents to areas which were not their primary residence prior to and during the lockdown period, which could slightly alter the data profile of local authorities. Our ecological study design serves as an important complement to analyses of individual-level patient data, to help understand how structural inequities at the population level affect COVID-19 burden. Here, we also adjusted for multiple potential confounders in our models to better identify associations between proportions of ethnicity groups in a local authority, or deprivation of the local authority, and COVID-19 case burden.

Our findings imply that structural inequities related to race/ethnicity, socioeconomic deprivation, local health disparities and environmental factors may be “root causes” of disparities in COVID-19 infection in England. The interaction between these factors is likely to multifaceted and complex [26], and may evolve over time. The identified clusters of COVID-19 caseload trajectory and their varied associations with contextual factors may also have important implications for future resource allocation strategies and public health planning, at the national level. Recent calls to the research community have highlighted the importance of reporting socioeconomic as well as racial/ethnic disparity data in order to properly understand COVID-19 health inequalities [26,35]. We conclude that public health practitioners and policymakers should recognize both the collective contribution of social determinants of health and their intersectionality, to better mitigate the burden of COVID-19 and reduce health disparities.

## Data Availability

All data is publicly available and provided by Public Health England.

https://www.gov.uk/coronavirus

## References

1 COVID-19 situation update worldwide, as of 18 May 2020. European Centre for Disease Prevention and Control. https://www.ecdc.europa.eu/en/geographical-distribution-2019-ncov-cases (accessed 18 May 2020).

2 Coronavirus (COVID-19) related deaths by ethnic group, England and Wales - Office for National Statistics. https://www.ons.gov.uk/peoplepopulationandcommunity/birthsdeathsandmarriages/deaths/articles/coronavirusrelateddeathsbyethnicgroupenglandandwales/2march2020to10april2020#ethnic-breakdown-of-deaths-by-age-and-sex (accessed 13 May 2020).

3 Aldridge RW, Lewer D, Katikireddi SV, et al. Black, Asian and Minority Ethnic groups in England are at increased risk of death from COVID-19: indirect standardisation of NHS mortality data. Wellcome Open Res 2020;5:88. doi:10.12688/wellcomeopenres.15922.1

4 Deaths involving COVID-19 by local area and socioeconomic deprivation - Office for National Statistics. https://www.ons.gov.uk/peoplepopulationandcommunity/birthsdeathsandmarriages/deaths/bulletins/deathsinvolvingcovid19bylocalareasanddeprivation/deathsoccurringbetween1marchand17april (accessed 18 May 2020).

5 Nicodemo C, Barzin S, Lasserson DS, et al. Population Vulnerability to Unexpected Health Shocks: Geographical Disparities in England at the Time of COVID-19. Rochester, NY: Social Science Research Network 2020. https://papers.ssrn.com/abstract=3571524 (accessed 13 May 2020).

6 Khunti K, Singh AK, Pareek M, et al. Is ethnicity linked to incidence or outcomes of covid-19? BMJ 2020;369. doi:10.1136/bmj.m1548

7 Bhala N, Curry G, Martineau AR, et al. Sharpening the global focus on ethnicity and race in the time of COVID-19. The Lancet 2020;0. doi:10.1016/S0140-6736(20)31102-8

8 Lancet T. Redefining vulnerability in the era of COVID-19. The Lancet 2020;395:1089. doi:10.1016/S0140-6736(20)30757-1

9 Coronavirus (COVID-19) in the UK. https://coronavirus.data.gov.uk/ (accessed 13 May 2020).

10 Population estimates - Office for National Statistics. https://www.ons.gov.uk/peoplepopulationandcommunity/populationandmigration/populationestimates (accessed 13 May 2020).

11 CRAN – Package dtwclust. https://cran.r-project.org/web/packages/dtwclust/index.html (accessed 13 May 2020).

12 Arbelaitz O, Gurrutxaga I, Muguerza J, et al. An extensive comparative study of cluster validity indices. Pattern Recognition 2013;46:243–56. doi:10.1016/j.patcog.2012.07.021

13 Fox S, Flowers J, Thelwall S, et al. fingertipsR: Fingertips Data for Public Health. 2020. https://CRAN.R-project.org/package=fingertipsR (accessed 13 May 2020).

14 English indices of deprivation 2019. GOV.UK. https://www.gov.uk/government/statistics/english-indices-of-deprivation-2019 (accessed 15 May 2020).

15 Standard Area Measurements (2019) for Administrative Areas in the United Kingdom. http://geoportal.statistics.gov.uk/datasets/standard-area-measurements-2019-for-administrative-areas-in-the-united-kingdom (accessed 13 May 2020).

16 Stier A, Berman M, Bettencourt L. COVID-19 Attack Rate Increases with City Size. Rochester, NY: Social Science Research Network 2020. https://papers.ssrn.com/abstract=3564464 (accessed 15 May 2020).

17 Iacobucci G. Covid-19: Deprived areas have the highest death rates in England and Wales. BMJ 2020;369. doi:10.1136/bmj.m1810

18 Barnett K, Mercer SW, Norbury M, et al. Epidemiology of multimorbidity and implications for health care, research, and medical education: a cross-sectional study. The Lancet 2012;380:37–43. doi:10.1016/S0140-6736(12)60240-2

19 Kontopantelis E, Mamas MA, van Marwijk H, et al. Chronic morbidity, deprivation and primary medical care spending in England in 2015-16: a cross-sectional spatial analysis. BMC Medicine 2018;16:19. doi:10.1186/s12916-017-0996-0

20 Baker C. Health inequalities: Income deprivation and north/south divides. Published Online First: 22 January 2019. https://commonslibrary.parliament.uk/insights/health-inequalities-income-deprivation-and-north-south-divides/ (accessed 15 May 2020).

21 Haynes Norrisa, Cooper Lisa A., Albert Michelle A. At the Heart of the Matter: Unmasking and Addressing COVID-19’s Toll on Diverse Populations. Circulation;0. doi:10.1161/CIRCULATIONAHA.120.048126

22 Dennison Himmelfarb CR, Baptiste D. Coronavirus Disease (COVID-19): Implications for Cardiovascular and Socially At-risk Populations. Journal of Cardiovascular Nursing 2020; **Publish Ahead of Print**. doi:10.1097/JCN.0000000000000710

23 Report 22 - Equity in response to the COVID-19 pandemic: an assessment of the direct and indirect impacts on disadvantaged and vulnerable populations in low- and lower middle-income countries. Imperial College London. http://www.imperial.ac.uk/medicine/departments/school-public-health/infectious-disease-epidemiology/mrc-global-infectious-disease-analysis/covid-19/report-22-equity/ (accessed 15 May 2020).

24 Walker P, Whittaker C, Watson O, et al. Report 12: The global impact of COVID-19 and strategies for mitigation and suppression. 2020. doi:10.25561/77735

25 Prats-Uribe A, Paredes R, Prieto-Alhambra D. Ethnicity, comorbidity, socioeconomic status, and their associations with COVID-19 infection in England: a cohort analysis of UK Biobank data. medRxiv 2020;:2020.05.06.20092676. doi:10.1101/2020.05.06.20092676

26 Chowkwanyun M, Reed AL. Racial Health Disparities and Covid-19 — Caution and Context. New England Journal of Medicine 2020;0:null. doi:10.1056/NEJMp2012910

27 Coronavirus (COVID-19) related deaths by occupation, England and Wales - Office for National Statistics. https://www.ons.gov.uk/peoplepopulationandcommunity/healthandsocialcare/causesofdeath/bulletins/coronaviruscovid19relateddeathsbyoccupationenglandandwales/deathsregistereduptoandincluding20april2020 (accessed 15 May 2020).

28 Employment by occupation. https://www.ethnicity-facts-figures.service.gov.uk/work-pay-and-benefits/employment/employment-by-occupation/latest (accessed 15 May 2020).

29 Geronimus AT, Hicken M, Keene D, et al. “Weathering” and Age Patterns of Allostatic Load Scores Among Blacks and Whites in the United States. Am J Public Health 2006;96:826–33. doi:10.2105/AJPH.2004.060749

30 Forde AT, Crookes DM, Suglia SF, et al. The weathering hypothesis as an explanation for racial disparities in health: a systematic review. Annals of Epidemiology 2019;33:1-18.e3. doi:10.1016/j.annepidem.2019.02.011

31 Krieger N. A glossary for social epidemiology. Journal of Epidemiology & Community Health 2001;55:693–700. doi:10.1136/jech.55.10.693

32 Devakumar D, Shannon G, Bhopal SS, et al. Racism and discrimination in COVID-19 responses. The Lancet 2020;395:1194. doi:10.1016/S0140-6736(20)30792-3

33 Medecins du Monde’s (MdM) 2019 Observatory Report Left Behind: The State of Universal Healthcare Coverage. MHADRI. 2019. https://mhadri.org/2019/12/17/medecins-dumondes-mdm-2019-observatory-report-left-behind-the-state-of-universal-healthcare-coverage/ (accessed 13 May 2020).

34 Oxford COVID-19 Impact Monitor. https://www.oxford-covid-19.com/ (accessed 18 May 2020).

35 Khalatbari-Soltani S, Cumming RG, Delpierre C, et al. Importance of collecting data on socioeconomic determinants from the early stage of the COVID-19 outbreak onwards. J Epidemiol Community Health Published Online First: 7 May 2020. doi:10.1136/jech-2020-214297

